# Investigating the clinical utility of biomarkers and other novel tests in younger onset neurocognitive disorders: the BeYOND study, protocol for a longitudinal clinical study in a real-world setting

**DOI:** 10.1101/2021.07.03.21259825

**Authors:** Samantha M Loi, Dhamidhu Eratne, Claire Cadwallader, Parsa Ravanfar, Carolyn Chadunow, Lesley Vidaurre, Sarah Farrand, Wendy Kelso, Anita MY Goh, Rosie Watson, Andrew Evans, Mark Walterfang, Dennis Velakoulis

**Affiliations:** Neuropsychiatry, Royal Melbourne Hospital, 300 Grattan Street, Parkville Australia 3050; Department of Psychiatry, University of Melbourne, Parkville 3052; Walter and Elizabeth Hall Institute of Medical Research, Parkville 3052; Florey Institute of Neuroscience and Mental Health, Parkville 3052; Melbourne Neuropsychiatry Centre, Department of Psychiatry, The University of Melbourne and Melbourne Health, Carlton South, VIC, Australia; National Ageing Research Institute, and Department of Psychiatry, University of Melbourne, Parkville 3052; Department of Medicine, Royal Melbourne Hospital, 300 Grattan Street, Parkville 3050

**Keywords:** neuropsychiatry, younger-onset dementia, longitudinal, neurodegenerative, diagnosis, biomarker

## Abstract

**Background:** Younger-onset dementia (YOD) can be challenging to diagnose due to its younger age of onset, heterogeneous aetiologies and broad range of presentations. Misdiagnosis is common with psychiatric conditions often diagnosed initially and diagnostic delay of five years is common. More information is needed to better understand and diagnose YOD, including the nature of symptom onset, progression of the disease, the relationship between cognition and functional outcomes for patients and carers, imaging changes and novel biomarkers. This paper reports on the background behind the “Investigating the clinical utility of biomarkers and other novel tests in younger-onset neurocognitive disorders”, the BeYOND study, and its methodology.

**Methods:** BeYOND is a clinically-oriented “real-world” longitudinal study that follows younger people presenting with an onset of neuropsychiatric symptoms ≤ 65 years of age. We aim to collect information on participants’ cognition, neuroimaging, mental health, and blood and cerebrospinal fluid (CSF) biomarkers at 18-month time-points over 3 years. We also aim to collect information regarding the experience of carers and/or family of participants.

**Conclusion:** Serial assessment of symptomatology, cognition, imaging, and blood and CSF biomarkers will be correlated with eventual diagnosis to determine the usefulness of these measures in determining a confident diagnosis. In addition, repeat measurements of the mental health and well-being of the participant and that of their carers/family while they traverse their diagnostic journey will provide important information about service provision and how they can be better supported.

## INTRODUCTION

Younger-onset dementia (YOD) represents approximately 9% of all dementias (Withall, 2013) and is defined as a dementia of which symptom onset occurs at 65 years or less. There are currently 28,000 people in Australia with this diagnosis (Dementia Australia statistics, 2021). YOD affects people who are usually in a productive stage of their lives in terms of employment and raising children, and can lead to significant psychosocial implications (Sansoni et al., 2016). Common causes of YOD include Alzheimer’s disease (AD) and frontotemporal dementia (FTD) (Loi et al., 2020a;Ferran et al., 1996). Genetic and metabolic causes of dementia include disorders such as Huntington’s disease and phenylketonuria are also associated with a younger age of onset (Kelley et al., 2008).

Previous work in the area has highlighted the various challenges related to YOD. These include the heterogeneous nature of symptom presentation which can lead to diagnostic delay (Draper et al., 2016;Loi et al., 2020b;van Vliet et al., 2013), difficulties with access to appropriate service provision for both the patient and their families (Sansoni et al., 2016;Cations et al., 2017), and the associated distress and psychiatric effects on both of these parties (van Vliet et al., 2011). In addition, authors have suggested diagnostic instability in YOD, particularly in people who present with psychiatric symptoms (Woolley et al., 2011;Perry et al., 2019). Diagnosis in this subgroup may change from a neurodegenerative condition to psychiatric condition and vice versa (Tsoukra et al. 2021 accepted). Currently, monitoring people with suspected YOD over time with repeat assessment is the commonest method to determine and provide accurate diagnosis. The uncertainty caused by a prolonged diagnostic delay can provoke significant ongoing distress for the symptomatic person, and for their carers due to potential consequences for future employment, financial stability, familial relationships, heritability and may also mean costly and potentially invasive investigations(van Vliet et al., 2011;van Vliet et al., 2013).

Novel biomarkers under current investigation have the potential to provide more efficient and accurate diagnoses. Neurofilament light chain (NfL) is a promising CSF biomarker reported by our group, to have 87% sensitivity and 90% specificity in distinguishing between psychiatric and neurodegenerative/neurological disorders (Eratne et al., 2020). The utility of blood plasma NfL is currently being investigated by our group. Less accurate odour identification may also provide some additional support for a diagnosis of a neurodegenerative disorder over a primary psychiatric disorder (Pachi et al., 2021).

Furthermore, quantitative susceptibility mapping (QSM) is an MRI technique which quantifies specific metals such as iron, copper and calcium, as well as blood degradation in areas of the brain, and in some disorders, is correlated with disease duration and severity (Ravanfar et al., 2021) Although, these novel biomarkers show promise, they are yet to be thoroughly validated. There is a clear need for high quality research studies to develop accurate, accessible and equitable diagnostic tools to diagnose YOD (Ducharme et al., 2020). This is likely to alleviate distress for the person with YOD and their families, and to provide information for future planning and service provision.

Longitudinal studies in YOD have provided important information regarding the aetiology, symptom presentation, and carer and patient perspective (Pasquier et al., 2004;van Vliet et al., 2010;Atkins et al., 2012). However, these studies have been limited in terms of follow-up duration, lack of neuroimaging and biomarker analysis, and lack of inclusion of a range of YOD conditions (Loi et al., 2020c). Furthermore, given that individuals with YOD may be misdiagnosed with a psychiatric condition due to the presence of psychiatric symptoms, it is arguably important to follow up and reassess these people in order to determine diagnostic stability as well as explore which investigations and assessments can provide more accurate, specific and sensitive information regarding diagnostic accuracy.

We present the protocol for BeYOND: Investigating the clinical utility of biomarkers and other novel tests in younger-onset neurocognitive disorders. This longitudinal cohort study aims to: 1) assess the frequency and aetiology of YOD; 2) correlate neuropsychiatric symptoms with cognitive measures; 3) investigate the clinical utility of novel biomarkers which may aid diagnosis such as CSF and plasma NfL; 4) assess diagnostic stability over time; and 5) review factors that contribute to carer and patient mental health and quality of life.

## METHODS

### Design

The BeYOND study is a longitudinal observational cohort study of people referred to Neuropsychiatry at the Royal Melbourne Hospital. Neuropsychiatry is a tertiary specialist centre located in metropolitan Melbourne (Victoria, Australia), and receives referrals for the investigation of a range of neuropsychiatric conditions, including YOD. As a public psychiatry service, common referrals include younger adults with chronic psychiatric conditions who have a suspected dementia.

### Study sample

We aim to recruit 120 individuals who are referred for a diagnostic assessment for possible YOD, over a two-year period. Inclusion criteria include onset of psychiatric, behavioural, neurological or cognitive symptoms prior to the age of 65 years who are referred for assessment for a possible YOD. Participants will have a minimum age limit of 18 years and will not have significant intellectual disability. A drop-out rate of 20% will be allowed for with regard to follow-up assessment data.

### Assessments

All participants will undergo thorough assessments within a multidisciplinary team, including neuropsychiatry, neurology, neuropsychology, and other assessments such as occupational therapy and speech pathology as required. The Neuropsychiatry Cognitive Assessment (NUCOG) will be used for bedside cognitive testing which assesses the five cognitive domains of attention, visuospatial, memory, executive and language (Walterfang et al., 2006). This 21-item tool provides a total score out of 100, and scores out of 20 for each of the five cognitive domains. The NUCOG has been shown to accurately differentiate between dementia, mild cognitive impairment and healthy controls. Consensus diagnostic criteria for dementia such as National Institute on Aging-Alzheimer’s Association (NIAA) (McKhann et al., 2011)for Alzheimer’s dementia, Rascovsky criteria for behavioural-variant frontotemporal dementia (FTD) (Rascovsky et al., 2011) and the Diagnostic Statistical Manual version V (DSM-V).

### Blood and CSF biomarkers (table 1)

As well as routine blood tests, blood samples will also be tested for NfL levels Blood serum and plasma will be collected, aliquoted, and stored at -80C. Plasma NfL will be analysed using ultrasensitive Quanterix Simoa technology. CSF collected in polypropylene tubes will be analysed for AB42, total tau, phosphorylated tau, using ELISA INNOTEST technology as previously described (Eratne et al., 2020), and from 2021 onwards using Roche Elecsys technology. CSF NfL will be measured using ELISA Umandiagnostics NF-Light kits as previously described (Eratne et al., 2020), and also using Quanterix Simoa technology. DNA material will be used to extract a range of unvalidated but known genetic risk markers for dementia (see Table 1).

**Table 1.** Routine blood tests and investigations

|  | **Routine** | **Additional** |
| --- | --- | --- |
| **Blood tests** | Full blood count, electrolytes, liver function tests, iron studies, fasting cholesterol and glucose, calcium, magnesium and phosphate, thyroid function tests, B12, folate<br>Autoimmune encephalitis blood screen * | Neurofilament light chain<br>Other research biomarkers such as neurogranin |
| **Neuroimaging** | Magnetic resonance imaging<br>SPECT<br>Positron emission tomography | Quantitative susceptibility mapping |
| **Cerebrospinal fluid (CSF)** | Lumbar puncture - $\alpha\beta$-42, phosphorylated tau and total-tau<br>Autoimmune encephalitis CSF screen * | Neurofilament light chain |
| **Genetic blood testing** |  | APOε4<br>Single gene testing such as C9orf72 or mutant huntingtin*<br>14-3-3*<br>Whole exome or whole genome sequencing* |
| **Depending on presentation** | ECG, EEG |  |

\*depending on clinical suspicion

### Neuroimaging

Participants will undergo neuroimaging as part of their standard clinical care. This includes structural magnetic resonance imaging (MRI). Sequences as part of the protocol include MPRAGE, T1 and T2 FLAIR, DWI and morphometry (axial, coronal and sagittal planes); single photon emission computed tomography (SPECT) and/or positron emission tomography (PET). Additionally, a subset of participants will be asked to undertake QSM imaging to investigate brain iron deposition. These will be participants who have a probable diagnosis of AD or FTD, based on CSF results or genetics.

### Neuropsychology

Comprehensive neuropsychological assessment will assist in determining diagnosis, disease severity and prognosis. Cognitive changes may precede neuroimaging changes, signifying early neurodegeneration. Domains assessed will include premorbid intelligence, attention and working memory, new learning and memory, speed of processing, speech and language, visuospatial and constructional skills and executive functioning. See Table 2 for details regarding specific neuropsychological tests.

**Table 2.** Neuropsychological assessments

|  |  |
| --- | --- |
| **BNT** | The Boston Naming Test (BNT) is a 60-item clinician-administered test measuring confrontational naming ability (Kaplan et al. 1983). It is widely used to assess anomia in clinical populations. |
| **CVLT-II** | The California Verbal Learning Test - second edition (CVLT-II) is a clinician-administered auditory verbal list-learning task that assesses new learning and memory (Delis et al. 2000). |
| **DKEFS** | The Delis-Kaplan Executive Function System (DKEFS) is a comprehensive battery of nine standardised subtests that assess executive functions (Delis et al. 2001). Subtests administered will include the Trail Making Test (mental switching), Colour Word Interference (self-monitoring/inhibition/divided attention and Verbal Fluency (phonemic and semantic verbal generativity). |
| **Rey-Osterrieth Complex Figure** | The Rey-Osterrieth Complex Figure (RCF) is a clinician-administered neuropsychological tool used to assess visuo-spatial perception/construction, perceptual organisation and memory (Rey & Osterrieth, 1993). The examinee is required to copy a complex line drawing made up of 18 components, with subsequent free-recall of the figure after three and 30 minutes. A score out of 36 (2 points for the inclusion and accuracy of each component) is calculated for each condition. A 24-item recognition task is also completed. |
| **Simple Copy** | Simple copy is a clinician-administered test that allows a qualitative assessment of basic visuospatial and visuoconstructional ability. The examinee is required to copy simple line drawings varying in complexity from a triangle to a bicycle. The test can be rated as intact, or the nature of difficulties can be described by the clinician. |
| **WAIS-IV** | The Wechsler Adult Intelligence Scale - Fourth Edition (WAIS-IV) is a comprehensive clinician-administered neuropsychological battery used to assess intellectual functioning (Wechsler, 2008). It includes 10 core subtests and five optional subtests that examine a wide range of cognitive abilities including information processing speed, attention and working memory and verbal and non-verbal skills. Core selected subtests will include Similarities (abstract verbal reasoning), Block |
|  | Design (visuo-constructional ability), Digit Span Forward and Backward (immediate auditory attention and working memory) and Coding (psychomotor speed). Optional subtests will be administered depending on clinical diagnostic utility. Optional tests include Vocabulary (word knowledge), Matrix Reasoning (visuospatial reasoning), Visual Puzzles (visuospatial reasoning and problem solving), Information (general knowledge), Arithmetic (mental arithmetic) and Symbol Search (psychomotor speed). |
| **WMS-IV** | The Wechsler Memory Scale, Fourth Edition (WMS-IV) is a comprehensive neuropsychological battery of memory functioning (Wechsler, 2009). It includes 10 subtests that assess memory across verbal and visual domains, including immediate and delayed memory, and working memory. Core subtests include Logical Memory I and II (verbal story memory and recall). Optional subtests include Visual Reproduction I and II (visual pattern memory and recall) and Verbal Paired Associates I and II (verbal new learning and recall). |
| **WTAR** | The Wechsler Test of Adult Reading (WTAR) is a clinician-administered word reading task used as a measure of premorbid intellectual functioning (i.e., intellectual functioning prior to the onset of a condition or injury affecting the brain) (Wechsler, 2001). Performance on this test is insensitive to the effects of a range of YOD conditions including mild Alzheimer's disease, Huntington's disease and Parkinson's Disease. The WTAR is co-normed with the Wechsler Adult Intelligence Scales, Third Edition. |

### Questionnaires for patients and carers

A range of questionnaire data will be collected from both the participants and their carers. These include questions about mental health and wellbeing, subjective cognitive complaints, ability to complete activities of daily living, personality and carer burden (Table 3).

**Table 3.**
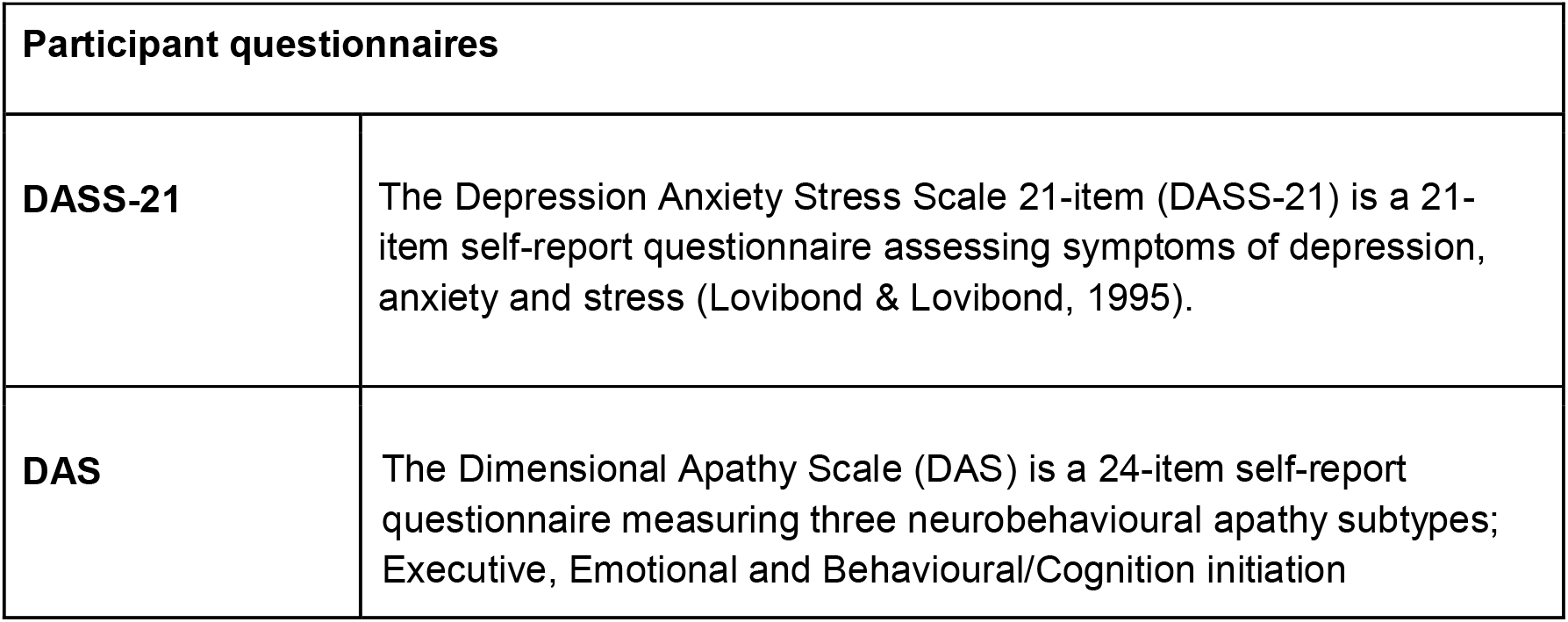
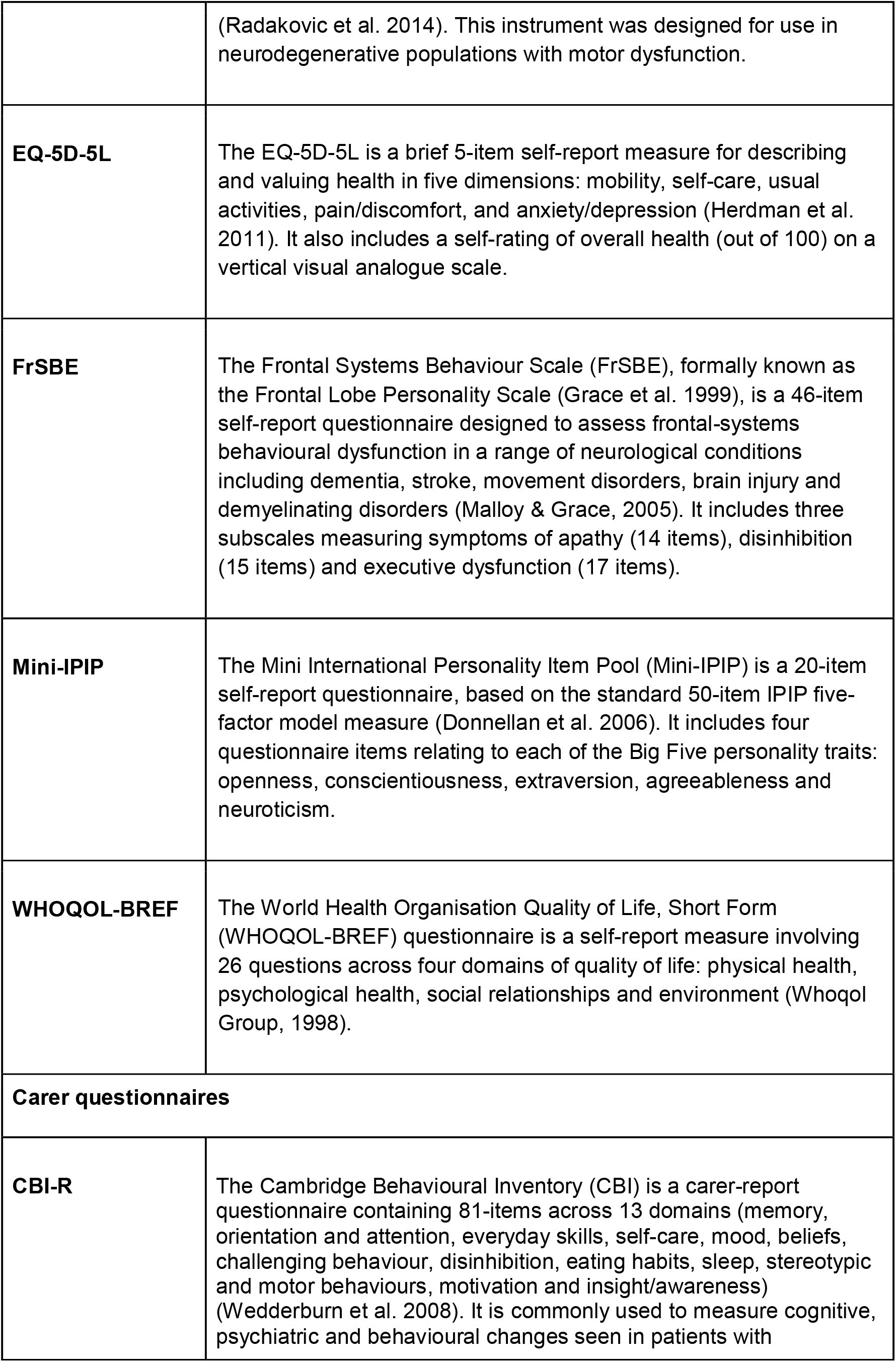
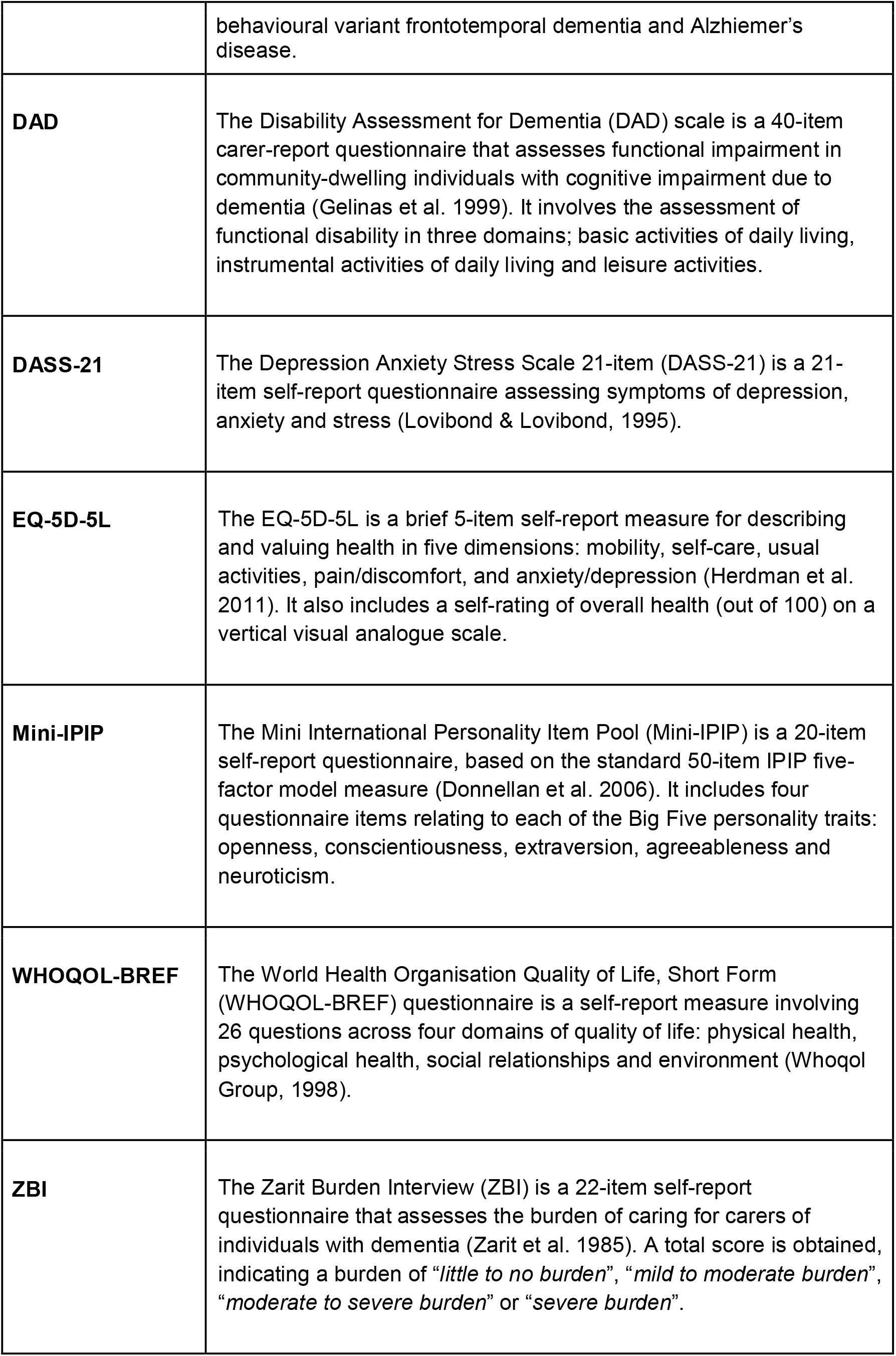
Participant and carer questionnaires

These assessments will be repeated 18 months after the baseline assessment where possible. Mortality information will be sought using the National Death Index.

### Statistical analyses

Data will be quantitative and two separate analyses will be conducted using the complete dataset and data from the full cohort with potential missing values. Categorical data such as gender and diagnosis will be analysed using categorical statistics e.g. Chi square and percentages. Continuous data will be checked for normality using skewness, kurtosis and Shapiro Wilks (or Kolmogorov-Smirnov) tests. For those variables which are normally distributed, parametric tests will be used. To compare variables from the different diagnostic groups t-tests and ANOVA will be used with Pearson correlations. For non-normal data, non-parametric tests such as Wilcoxon and Spearman correlations will be used. For comparing NfL between diagnostic groups, GLM with age as a covariate (given age strongly correlates with NfL levels). For analyses of repeated measures, a mixed effect modelling approach will be implemented within the Bayesian framework which will allow incorporation of prior scientific information about the diagnostic tests into the modelling process. This will result in reliable model estimates incorporating uncertainty in parameter estimation. Depending on the type of analyses, either R or SPSS will be used to analyse the data.

### Ethical considerations

This project was approved by the local hospital human research ethics committee (MH 2018.371).

## DISCUSSION

BeYOND will be the first longitudinal study to investigate YOD in a clinical “real-world” setting. This is achieved through the inclusion of younger adults presenting with a range of neuropsychiatric symptoms, referred to our service for diagnostic investigation of YOD. This reflects the “real-world” practice and in contrast to a well-characterised dementia cohort, defined by a narrow set of symptoms or diagnosis after the fact. The BeYOND methodology reflects the AIBL study which includes well-characterised people with AD, and also includes people with mild cognitive impairment and subjective memory complaints, with the aim of following up these people with “unstable” diagnoses and categorising them over time. Similarly, BeYOND recognises that diagnoses are “unstable” and may change over time. By repeat assessments of neuroimaging and biomarkers, we will be able to provide much needed information about the relationships between these markers, neuropsychiatric and cognitive symptoms which will help predict who might “convert” to a dementia or who may not. This will help contribute to decrease the diagnostic delay and provide more accurate diagnoses earlier in the assessment journey.

Limitations include the specialisation of our public psychiatry service which may create a sample bias regarding the nature of patients referred for diagnostic clarification of YOD. For example, there is likely to be bias towards referrals for people with chronic psychiatric disorders and behavioural problems (ie. less language-related and/or motor dysfunction symptom presentations of YOD). While we are a state-wide service and not restricted to community dwelling people, due to the need to attend the service, those in residential accommodation or from rural and remote locations may be under-represented in the study sample.

This study will provide important information on YOD for patients and families regarding their diagnostic experiences and post-diagnostic care. This information will be used to improve clinical service provision and inform future models of care.

## Data Availability

Not applicable currently

